# How Much Time Do Surgeons Spend Operating?

**DOI:** 10.1101/2025.02.15.25321677

**Authors:** Nicholas S. Andrade

**Affiliations:** Department of Neurosurgery, Christus Mother Frances Hospital, Tyler, TX 75701, USA

## Abstract

**Background:** Although surgeons participate in various clinical activities, performing surgery is their unique specialized function. Despite this, the fundamental metric of time spent operating remains unexplored.

**Methods:** We retrospectively analyzed operating room cases performed by surgeons who worked full-time for a full calendar year in two general hospitals. The operating time (from start to stop of the procedure) was extracted from electronic medical records for all cases from 2021 to 2024.

**Results:** 263 surgeon-years were analyzed across 14 specialties. Surgeons spent a median of 451 hours operating annually (range 52-1160 hours) and performed a median of 414 cases (49-1381 cases). Annual operating time varied tenfold across specialties, from Cardiothoracic surgery (856 hours) to Podiatry (87 hours).

**Conclusion:** Surgeons spend about a quarter of their work time performing their uniquely trained function. The substantial variation within and between specialties suggests opportunities to optimize surgical workforce utilization. Understanding the determinants of operating time and sustainable surgical workload could improve healthcare system efficiency and surgical care delivery.

---

Division of labor and specialization have driven modern productivity gains across industries, yet their application to surgical practice remains complex. Surgeons represent a uniquely specialized workforce whose technical expertise requires 5-7 years of dedicated training. Recent studies predict continued surgical workforce shortages through 2050 (1; 2; 3), yet we lack fundamental data about how much time surgeons spend performing their specialized function—operating.

Operating time represents a crucial metric for individual surgeons and healthcare systems. Higher surgical volumes correlate with better patient outcomes(4; 5), while increased operating time could help address workforce shortages(6) and reduce burnout(7; 8; 9), as most surgeons chose their specialty primarily to perform surgery. However, the modern surgeon’s workday includes multiple responsibilities, including clinic visits, inpatient rounds, administrative duties, and teaching. While documentation burdens have increased, the expansion of advanced practice providers (APPs) offers opportunities to redistribute non-operative tasks.

Existing studies of surgical productivity typically focus on output measures such as case volumes or relative value units (RVUs)(10), overlooking time as a critical constraining resource. This analysis aims to establish the first dataset of annual operating time per surgeon across different specialties, evaluate variations in operating patterns, and identify opportunities to better leverage surgeons’ specialized skills.

## 1 Methods

We conducted a retrospective cohort study that analyzed operating room cases from two general hospitals in Tyler, TX, USA, from January 1, 2021, to December 31, 2024. Both hospitals are acute care, tertiary referral facilities with 400-500 beds performing approximately 15,000 surgical procedures annually. Data were extracted from the electronic medical record system. All surgical specialties were included in the analysis. For each case, we collected the following variables: surgeon identifier, surgical specialty, procedure start time (defined as initial incision), and procedure end time (defined as final closure). No personal health information was collected; therefore, IRB approval was not required. Only surgeons with full-time status and complete calendar year data were included in the analysis. A surgeon-year was defined as the caseload of a particular surgeon who worked full-time during a complete calendar year, to measure steady-state productivity and eliminate the effects of workforce turnover. Statistical significance was assessed using two-tailed z-tests comparing each specialty’s mean to the overall population mean, with standard errors calculated using specialty-specific standard deviations. Differences were considered statistically significant at p<0.05. Cases with missing time stamps or inconsistent data entries were excluded from the analysis (<1% of total cases). Data processing and analysis were performed using Python in Google Colab. Manuscript preparation and statistical analysis review were assisted by Claude (Anthropic, 2024), an artificial intelligence language model, with all outputs validated by human review and final decisions made by the authors.

## 2 Results

Analysis included 263 surgeon-years across 14 surgical specialties. With respect to operating hours, several specialties showed statistically significant deviations from the mean. Cardiothoracic surgery and Surgical Oncology had significantly higher operating hours, while Podiatry, Ophthalmology, Gynecology, and Trauma had significantly lower operating hours. Average case duration showed significant variation across specialties. Cardiothoracic (148 minutes), Surgical Oncology (121 minutes), and Plastic surgery (106 minutes) had significantly longer case lengths. Conversely, Ophthalmology (24 minutes) and Podiatry (32 minutes) had significantly shorter case durations. All specialties except ENT showed statistically significant differences in case length from the mean. Regarding annual case volume, Neurosurgery and Ophthalmology (616 cases) performed significantly more cases than the mean, while Trauma (121 cases), Podiatry (163 cases), Gynecology (194 cases), and Gyn Onc (284 cases) performed significantly fewer cases. Orthopedic surgery (520 cases) and Plastic surgery (333 cases) also showed statistically significant differences in case volume.

## 3 Conclusions

Analysis of surgeon operating time across multiple specialties has revealed that surgeons spend approximately 22% of a standard work year (median 451 hours annually) performing surgery. This utilization varies dramatically across specialties, ranging from 856 hours annually for Cardiothoracic surgery to 87 hours for Podiatry. These variations reflect fundamental differences in practice patterns, case complexity, and non-operative responsibilities.

The contrast between specialties is apparent in their case volumes and durations. Some specialties, like Ophthalmology, perform many short procedures (616 cases/year, averaging 24 minutes), while others, such as Cardiothoracic surgery, conduct fewer but longer operations (351 cases/year, averaging 148 minutes). Certain specialties, including Trauma Surgery, maintain significant non-operative critical care responsibilities, which naturally limit their dedicated operating room time. Many specialties are a mixture of medical management, office procedures, and surgical procedures.

The surgical process has evolved to incorporate numerous specialized roles that improve efficiency and quality of care.(11) An outpatient surgical patient can be cared for by more than a dozen care providers. Some service lines have developed even more specialized models, such as transplant surgery’s use of dedicated medical specialists for comprehensive patient care. Despite these developments in care team models, the relatively low proportion of time surgeons spend operating suggests opportunities for further optimization.

Surgeon workforce shortages are one of several challenges facing healthcare systems that suggest the need for innovative approaches to surgical practice.(1; 2) One potential direction is the separation of surgical decision-making from technical operative delivery. This model could allow surgeons to focus on technical operative care while dedicated diagnosticians manage patient evaluation and procedural selection. Such specialization could optimize for different skill sets: clinical judgment and communication abilities for diagnosticians, and technical excellence and operative efficiency for proceduralists. The current model assumes that surgeons will excel at all of the above.

Future research should examine the determinants of operating time variation, the impact of advanced practice providers, the effectiveness of alternative practice models, and the relationship between operating time and both patient outcomes and surgeon satisfaction. The current study is limited–it involves only two hospitals in a specific region, and findings may differ in a broader study. However, we suggest that surgeon operating time is a fundamental metric worthy of further study, for the individual surgeon’s time management and for the healthcare system at large.

**Table 1:** Operating Hours by Specialty.

| Specialty | Mean | p-value |
| --- | --- | --- |
| Cardiothoracic | 856 | <0.01 |
| Surgical Oncology | 822 | <0.01 |
| Neurosurgery | 574 | 0.15 |
| Plastic | 557 | 0.11 |
| General | 555 | <0.01 |
| Orthopedic | 496 | 0.12 |
| Urology | 454 | 0.99 |
| Gyn Onc | 451 | 0.97 |
| Vascular | 408 | 0.34 |
| ENT | 394 | 0.42 |
| Ophthalmology | 196 | <0.01 |
| Gynecology | 194 | <0.01 |
| Trauma | 154 | <0.01 |
| Podiatry | 87 | <0.01 |

**Figure 1:**
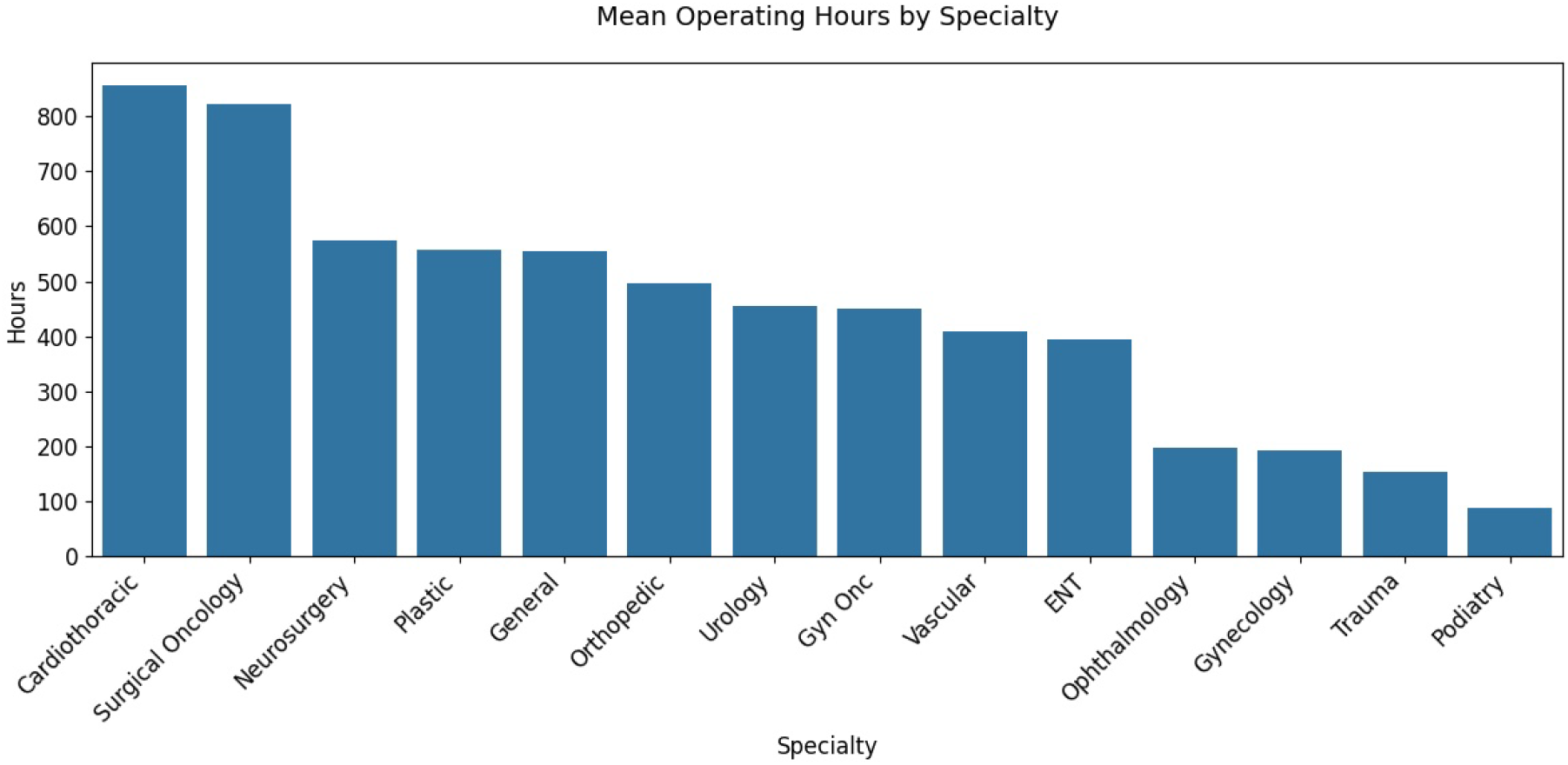
Mean Operating Hours by Specialty.

**Table 2:** Case Length by Specialty (Minutes)

| Specialty | Mean | p-value |
| --- | --- | --- |
| Cardiothoracic | 148 | <0.01 |
| Surgical Oncology | 121 | <0.01 |
| Plastic | 106 | <0.01 |
| Gyn Onc | 95 | 0.06 |
| General | 81 | 0.03 |
| Trauma | 76 | 0.48 |
| Urology | 64 | 0.27 |
| Orthopedic | 61 | 0.01 |
| Gynecology | 60 | 0.34 |
| Vascular | 57 | 0.04 |
| Neurosurgery | 56 | 0.18 |
| ENT | 49 | 0.03 |
| Podiatry | 32 | <0.01 |
| Ophthalmology | 24 | <0.01 |

**Figure 2:**
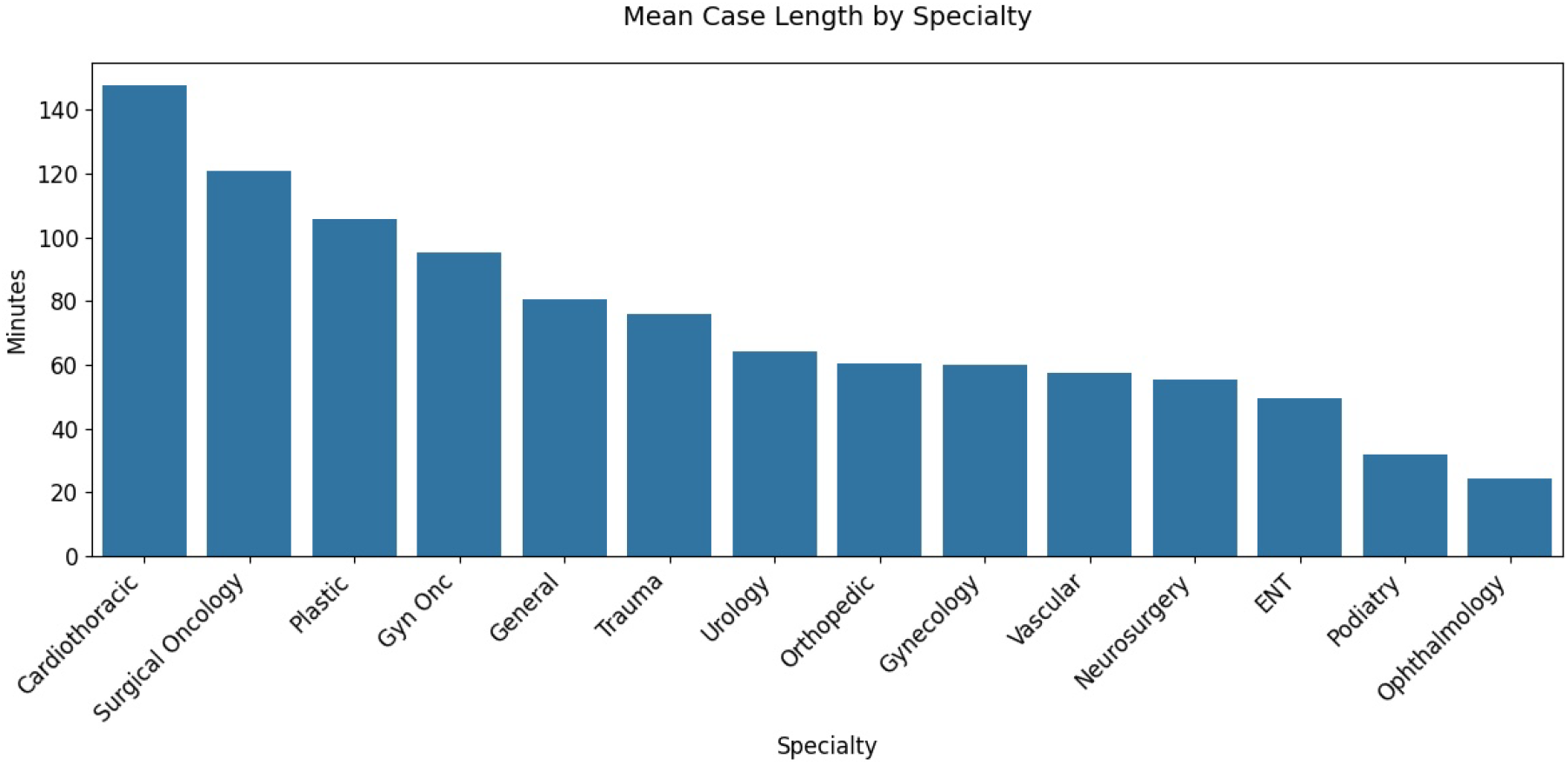
Mean Case Lengths by Specialty.

**Table 3:** Number of Cases by Specialty.

| Specialty | Mean | p-value |
| --- | --- | --- |
| Neurosurgery | 622 | <0.01 |
| Ophthalmology | 616 | <0.01 |
| Orthopedic | 520 | <0.01 |
| ENT | 460 | 0.46 |
| General | 454 | 0.18 |
| Vascular | 435 | 0.62 |
| Urology | 428 | 0.71 |
| Surgical Oncology | 417 | 0.96 |
| Cardiothoracic | 351 | 0.24 |
| Plastic | 333 | 0.14 |
| Gyn Onc | 284 | 0.12 |
| Gynecology | 194 | <0.01 |
| Podiatry | 163 | <0.01 |
| Trauma | 121 | <0.01 |

**Figure 3:**
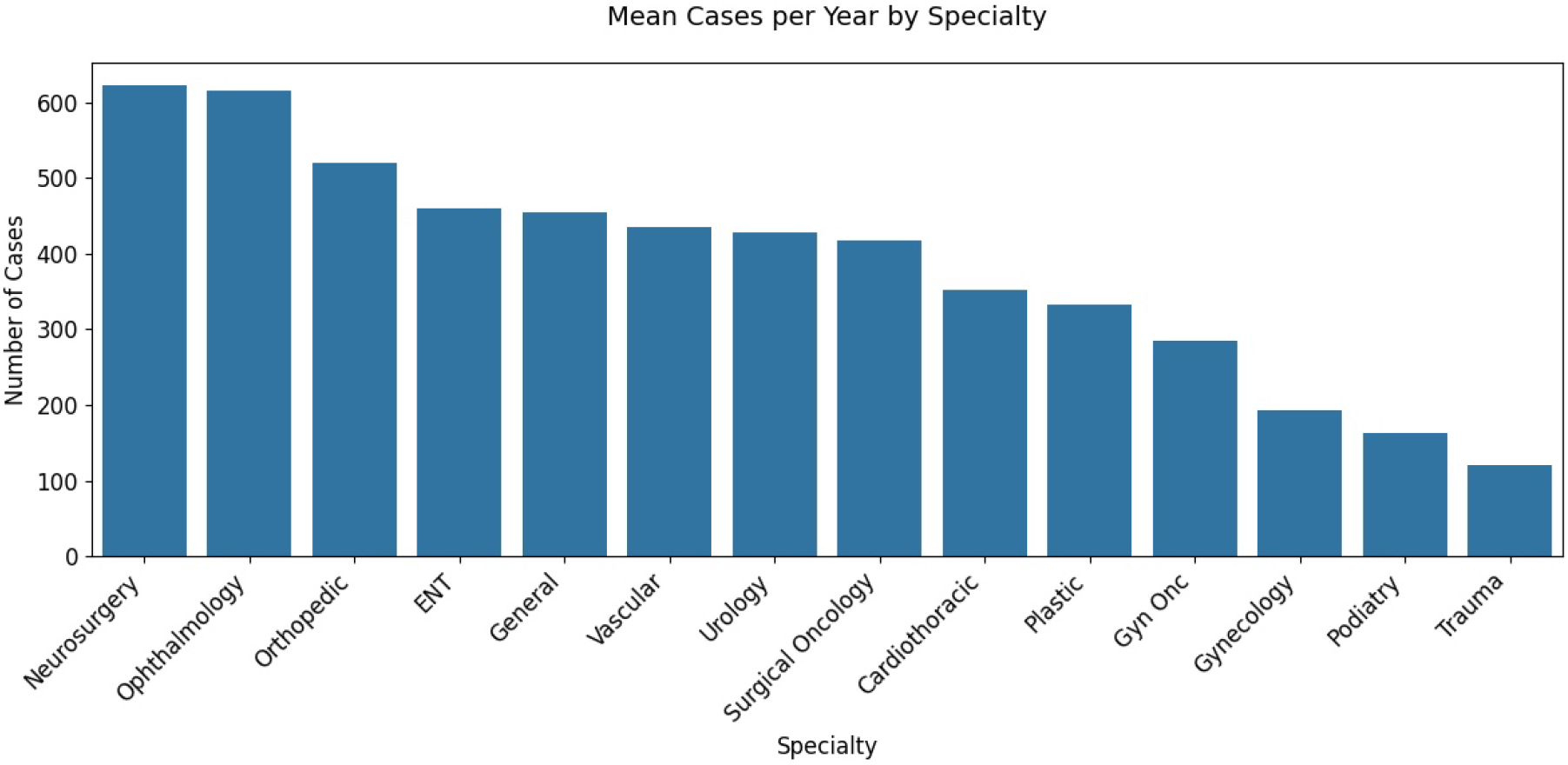
Mean Cases per Year by Specialty.

**Figure 4:**
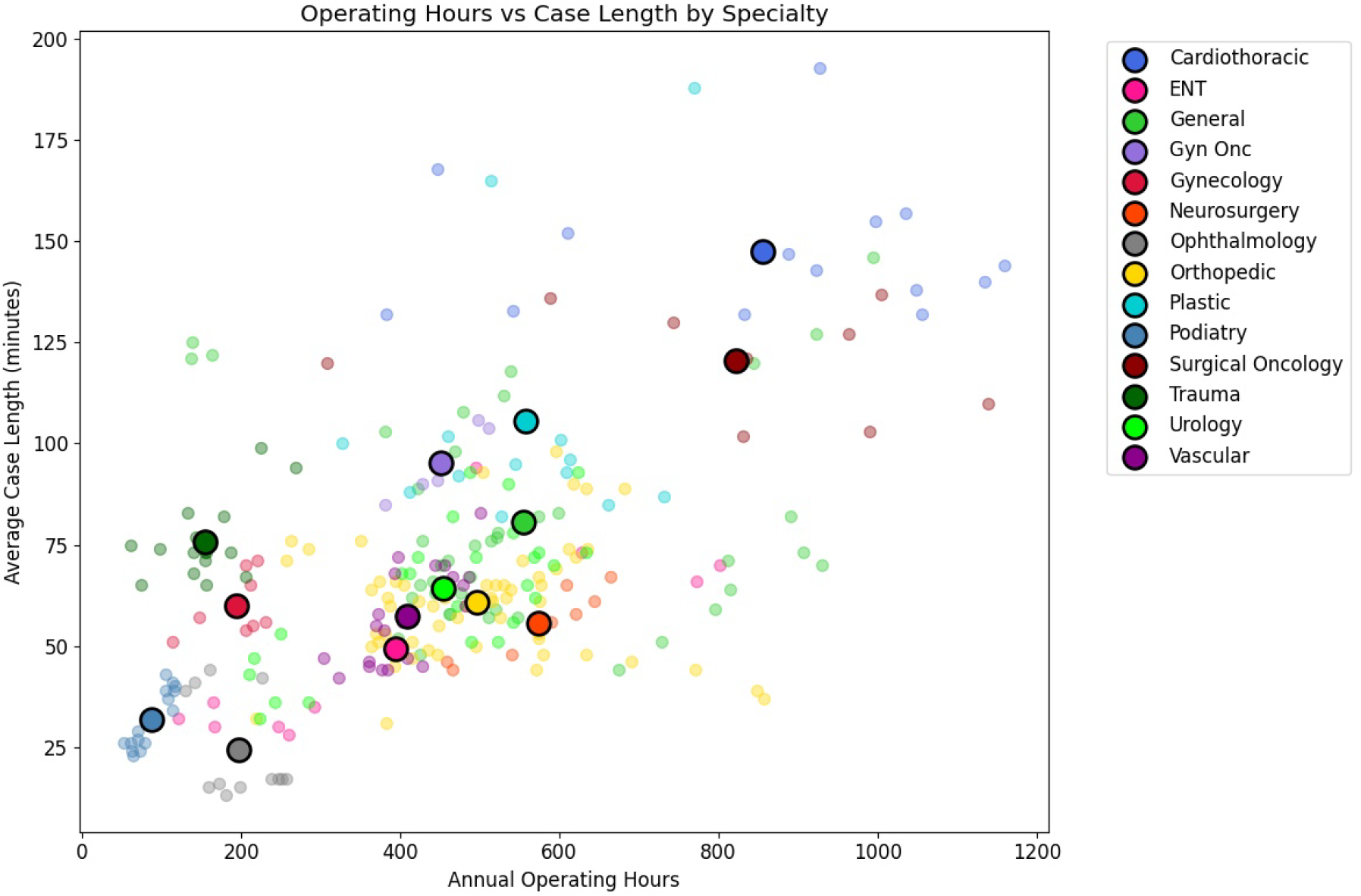
Scatter plot of annual operating hours versus average case length by specialty. Small dots represent individual surgeon-years, while large dots with black edges show specialty averages.

## Data Availability

All data produced are available online at Github

https://github.com/nsa122/surgeon-hours

## Acknowledgments

Thanks to Mateo Ziu, M.D., Shane Hawksworth, M.D., and Mitchell George, M.D. for their helpful comments.

## Author contributions

N.S.A. conceived the study design, did the analysis, and wrote the paper.

## Data and materials availability

Code and data are available at https://github.com/nsa122/surgeon-hours

